# Modelling maternal cytomegalovirus seroprevalence in Australia using maternal country of birth: Implications for antenatal screening

**DOI:** 10.1101/2025.08.21.25333872

**Authors:** Melvin Barrientos Marzan, Tanya Tripathi, Liv Dumville, Jo Watson, Natasha E Holmes, Lisa Hui

## Abstract

**Background:** Cytomegalovirus (CMV) is the most common congenital infection and a leading preventable cause of neurodevelopmental disability. Contemporary maternal CMV seroprevalence estimates are needed to inform antenatal screening policy in Australia.

**Purpose:** To estimate CMV seroprevalence among women referred for antenatal care at a tertiary maternity hospital, identify clinical and sociodemographic associations with serostatus, and model national and hospital-level seroprevalence by maternal country of birth.

**Materials and Methods:** CMV serology results from GP antenatal referrals to a Melbourne tertiary hospital over 13 months were analysed. Seroprevalence was stratified by maternal country of birth, grouped by Organisation for Economic Co-operation and Development (OECD) membership. National and hospital-specific seroprevalence were modelled using Australian Bureau of Statistics and research dataset country-of-birth distributions.

**Results:** Of 4377 referrals, 591 (12.5%) included CMV serology; 61.3% were CMV IgG positive. Median gestational age at testing was 5.4 weeks; referral occurred at a median of 9 weeks. Seroprevalence was higher among women born in non-OECD countries (86.6%) versus OECD countries (54.3%) (adjusted OR 6.44, P<0.001). Modelled national maternal seroprevalence was 63.6%. Across Melbourne public hospitals, estimates ranged from 60% to 74%, reflecting demographic variation. Among 328 women tested for CMV IgM, 17 (5.2%) had positive or equivocal results; one had low avidity IgG, consistent with recent primary infection.

**Conclusions:** Maternal CMV seroprevalence in this cohort is higher than previously reported and strongly associated with maternal country of birth. These data inform evaluation of antenatal CMV screening feasibility and cost-effectiveness studies in Australia.

## Introduction

Cytomegalovirus (CMV) is the most common congenital infection, present in 1 in 200 births, and a leading preventable cause of neurodevelopmental impairment and sensorineural hearing loss in high-income countries [1]. Primary maternal CMV infection in the first trimester carries the highest risk of fetal transmission and severe sequelae, with vertical transmission occurring in approximately 37% of cases and adverse outcomes in up to 19% of infected fetuses [2].

Despite this, CMV remains under-recognised in antenatal care. In Australia, the recently updated national Pregnancy Care Guidelines recommend targeted CMV IgG testing for pregnant individuals at increased risk of infection, namely, those with young children in the home, and childcare workers [3]. Growing evidence demonstrating the effectiveness of maternal valaciclovir therapy in reducing vertical transmission has sparked interest in universal antenatal CMV screening [4], but this data was not addressed in the updated Pregnancy Care Guidelines.

A global meta-analysis by Ssentongo et al. (2021) demonstrated that prevalence and burden of cCMV varies significantly according to national income classification [1]. Accurate, contemporary data on maternal CMV seroprevalence are essential for evaluating the feasibility and cost-effectiveness of targeted and universal screening approaches. Australian studies on maternal CMV seroprevalence are limited, and more than 20 years old. A single-centre prospective study from the eastern suburbs of Sydney in 2005 reported a seroprevalence of 57% [5]. More recently, our group published the results of small audit of 115 pregnancies with CMV serology performed at the booking visit, demonstrating that maternal country of birth was a stronger predictor of CMV serostatus than socioeconomic status, parity or other maternal characteristics [6].

Given Australia’s multicultural population, understanding the relationship between maternal country of birth and CMV serostatus is vital for informing risk-based screening strategies. The aim of this study was to generate updated estimates of maternal CMV seroprevalence using real-world antenatal data from a diverse Australian maternity population and to model local and national seroprevalence based on maternal country of birth. This evidence will help determine the potential value and impact of antenatal CMV screening within the Australian healthcare context.

The specific objectives of this study were: (i) to estimate CMV seroprevalence among women referred for antenatal care at [redacted - hospital name]; (ii) to assess the relationship between CMV seroprevalence and maternal sociodemographic factors; and (iii) to model local and national CMV seroprevalence based on the income classification of maternal country of birth.

## Methods

This study was conducted as part of a project on the feasibility and acceptability of antenatal CMV screening (Mercy Health HREC approval 2023-019, protocol v4.0, 04 December 2024). Ethical approval for use of 2023 data from the birth records dataset was obtained under approved data-sharing provisions (Austin Health HREC 64722 and Mercy Health HREC 2020-031).

Consecutive GP referrals for antenatal care at the XXXX were prospectively screened for CMV serology results between December 2023 and December 2024. Although the hospital does not recommend CMV serology as part of standard antenatal investigations, this testing is known to be initiated by GPs in approximately 13% of referrals [6]. Any referral containing CMV serology as part of the initial antenatal investigations was included in this analysis. Our prior audit of GP screening practices showed that women who had a child aged less than five years of age were not more likely to be screened for CMV by their GP, suggesting non-selective screening practices by our referring GPs [6]. Women who underwent CMV testing within the hospital due to clinical indications (e.g. fetal anomalies or maternal symptoms) were excluded.

The results of CMV serology (including CMV IgG, and IgM and IgG avidity testing if performed) were manually extracted and entered into REDCap (Research Electronic Data Capture) hosted at [XXX ] (ref). Each entry was checked by a second researcher for accuracy. Individual-level data included gestational age at testing and referral for antenatal care, maternal age, parity, and Social and Economic Index for Areas (SEIFA) based on residential postcode. SEIFA–IRSAD scores from the 2020 Australian Census were used to assess area-level socioeconomic status [7]. The original 1–10 scale, where 1 indicates the most disadvantaged and 10 the most advantaged areas, was recoded into tertiles for analysis. Group ‘1’ (most disadvantaged) comprised scores 1-6, Group ‘2’ comprised scores 7 and 8, and Group ‘3’ (most advantage) comprised scores 9 and 10. [7]. Maternal country of birth was categorised as either OECD or non-OECD according to the 2024 OECD membership list [8].

To estimate national CMV seroprevalence, we applied a weighted average of the OECD and non-OECD seroprevalence rates using national demographic data from the Australian Bureau of Statistics (2021). [9]. We further applied this model to estimate maternal CMV seroprevalence across public maternity services in Melbourne. Maternal country of birth data was obtained via secondary analysis of a research dataset that contained birth records from all Melbourne public hospitals [10].

Statistical analyses were performed using Stata version 19 [11]. Descriptive statistics were used to summarise participant characteristics and the distribution of cytomegalovirus (CMV) serostatus across key sociodemographic groups. Chi-squared tests and t-tests (or non-parametric equivalents, where appropriate) were conducted to assess univariate associations between covariates and CMV seropositivity. Subsequently, multivariable logistic regression models were fitted to estimate adjusted odds ratios (aORs) and 95% confidence intervals (CIs) for predictors of CMV seropositivity, including variables such as age, sex, country of birth, socioeconomic status, and other relevant demographic indicators. Variables were selected a priori based on existing literature and subject-matter expertise.

## Results

During the 13-month study period, 4733 consecutive referrals for antenatal care were screened, of which 591 (12.5%) contained CMV serology results. Of the 591 with CMV serology, the mean maternal age was 33.9 years and 44.3% were nulliparous. The median gestational age at CMV serology was 5.6 weeks (IQR: 4.6–7.1), while the referral for antenatal care occurred at a median of 8.4 weeks (IQR: 6.9– 10.1) (Figure 1). Most pregnant women (93.7%, 554/591) who were screened for CMV had their testing ≤ 12 weeks’ gestation; 21.5% were born in a non-OECD country.

**Figure 1.**
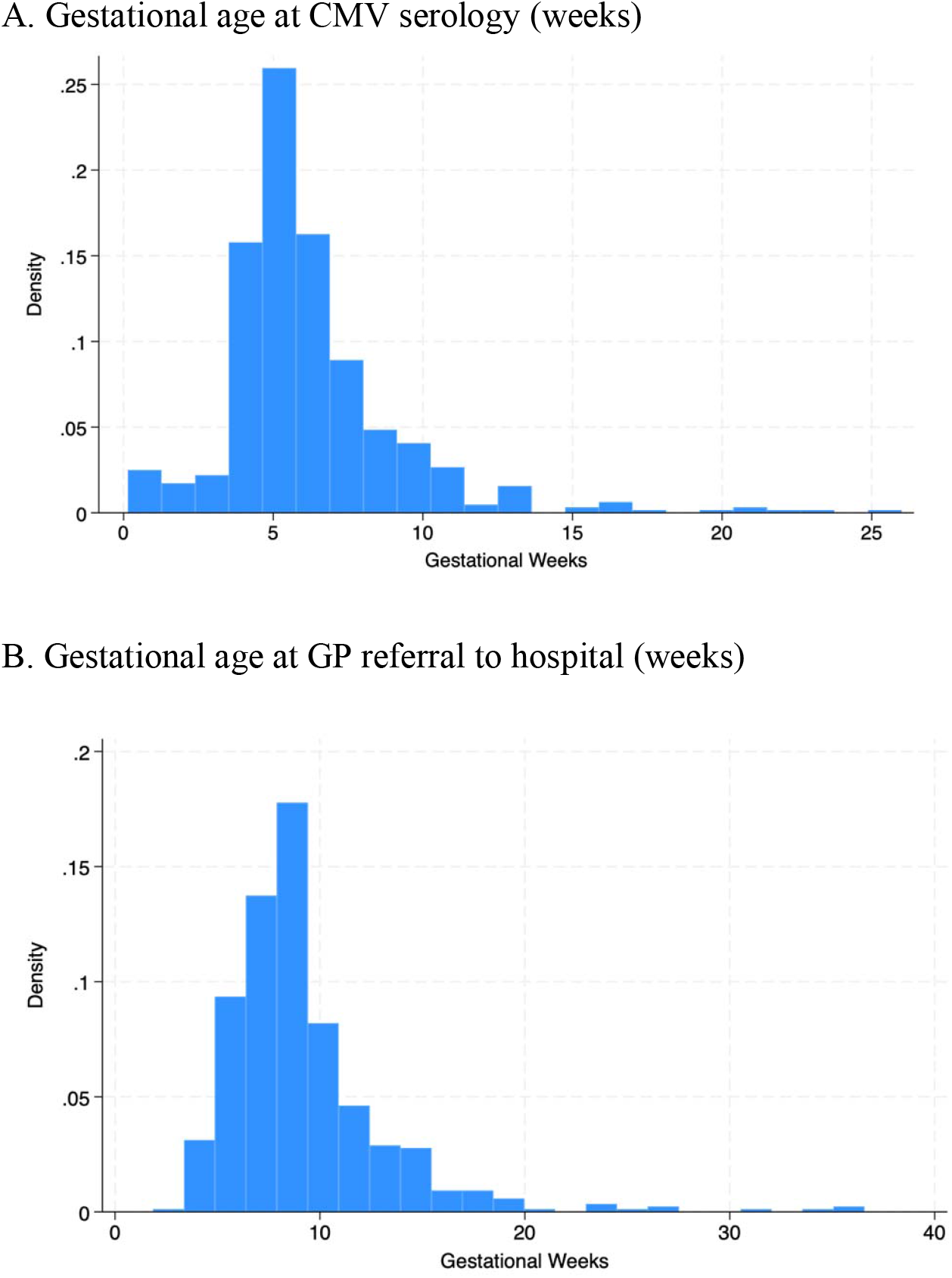
Gestational age distribution at CMV serology and GP referral to hospital

### *CMV seroprevalence at the XXXX* hospital

Of the 591 screened women, 362 (61.3%, 95% CI 57.3–65.2) were CMV IgG positive, and 229 (38.7%, 95% CI 34.8–42.7) were CMV IgG negative. The characteristics of the cohort by CMV serostatus are provided in Table 1.

**Table 1.** Characteristics of CMV seronegative (IgG−) and seropositive (IgG+) women at the XXXX.

|  | *Total CMV*<br>*screened*<br>*cohort* | *IgG (-)* | *IgG (+)* | *P-value* |
| --- | --- | --- | --- | --- |
| *Count (%)* | 591 | 229 (38.7) | 362 (61.3) |  |
| **Age at conception in Years, mean (SD)** | 33.9 (4.4) | 33.19<br>(4.2) | 33.50<br>(4.50) | 0.41 |
| **Maternal age category, years (%)** |  |  |  |  |
| <29 | 114 | 50 (43.9) | 64 (56.1) | 0.33 |
| 30-34 | 265 | 95 (35.9) | 170 (64.2) |  |
| ≥ 35 | 118 | 76 (39.2) | 118 (60.8) |  |
| **OECD classification for country of birth, n (%)** |  |  |  |  |
| Non-OECD countries | 127 | 17 (13.4) | 110 (86.6) | <0.001 |
| OECD countries* | 464 | 212 (45.7) | 252 (54.3) |  |
| **Aboriginal Status (%)** |  |  |  |  |
| Not First Nations | 570 | 222 (39.0) | 348 (61.1) | 0.46 |
| First Nations | - | <5 | <5 |  |
| Not stated | - | <5 | <5 |  |
| **Parity, n (%)** |  |  |  |  |
| 0 | 262 | 111 (42.4) | 151 (57.6) | 0.11 |
| ≥1 | 329 | 112 (35.7) | 211 (64.1) |  |
| **SEIFA quintile, n (%)** |  |  |  |  |
| 1 - Most disadvantaged | 246 | 79 (32.1) | 167 (67.9) | 0.01 |
| 2 | 137 | 56 (40.9) | 81 (59.1) |  |
| 3 - Most advantaged | 196 | 90 (45.9) | 106 (54.1) |  |
| **Referral date (%)** |  |  |  |  |
| ≤12 gestation weeks | 495 | 201 (40.6) | 294 (59.4) | 0.04 |
| ≥12 gestation weeks | 96 | 28 (29.2) | 68 (70.8) |  |
| **Testing date (%)** |  |  |  |  |
| ≤12 gestation weeks | 554 | 215 (38.8) | 339 (61.2) | 0.91 |
| ≥12 gestation weeks | 37 | 14 (37.8) | 23 (62.2) |  |

OECD, Organization of Economic Cooperation and Development.
\*OECD Countries - Australia, Austria, Belgium, Canada, Chile, Colombia, Costa Rica, the Czech Republic, Denmark, Estonia, Finland, France, Germany, Greece, Hungary, Iceland, Ireland, Israel, Italy, Japan, South Korea, Latvia, Lithuania, Luxembourg, Mexico, the Netherlands, New Zealand, Norway, Poland, Portugal, the Slovak Republic, Slovenia, Spain, Sweden, Switzerland, Turkey, the United Kingdom, and the United States

The factor most strongly associated with CMV serostatus was OECD status of maternal country of birth (Table 2). The seroprevalence among women born in non-OECD countries was 86.6% (110/127), significantly higher than women born in an OECD country (54.3%, 252/464, p<0.001). The adjusted OR of CMV seronegative status for women born in an OECD country was 6.44; (95% CI 3.42-12.13).

**Table 2.**
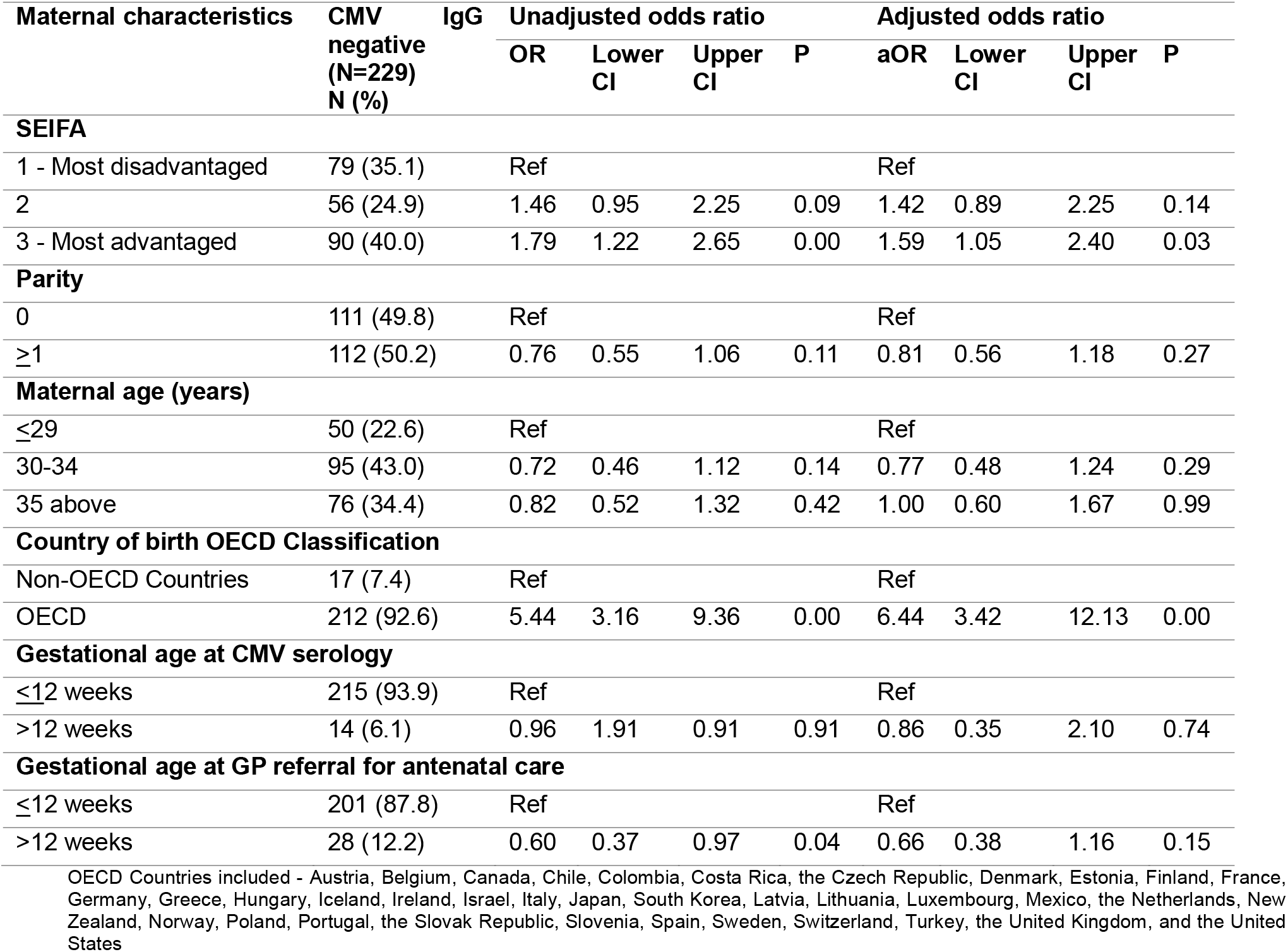
Unadjusted and adjusted odds ratio of maternal CMV seronegative status.

Residing in the most advantaged postcodes was also associated with a higher probability of being CMV seronegative compared with the least advantage postcodes. (aOR 1.69, 95% 1.05-2.40). Compared with seropositive women, a higher proportion of seronegative women were referred to the hospital for antenatal care before 12 weeks’ gestation. However, this association was not significant after adjusting for SEIFA, parity, maternal age, OECD country of birth, and gestation at serology (Table 2).

### CMV IgM and IgG avidity results

Of the 591 with CMV IgG serology, 328 (55.5%) had concurrent IgM testing reported. Of these, 17 (5.2%, 95% CI 2.8–7.6) had positive (n=15) or equivocal (n=2) CMV IgM results. CMV IgG avidity testing was performed on 13/15 with positive CMV IgM. Nine had high avidity, indicating infection at least 3 months prior to testing (i.e. preconception infections), 3 had moderate avidity (undetermined timing of infection), and 1 had low avidity (infection within 3 months of testing).

### Modelled national and local seroprevalence

According to ABS data, 27.51% of people giving birth in Australia were born in non-OECD countries and 72.49% were born in Australia or other OECD countries. The modelled national maternal CMV seroprevalence using the formula (0.2751 x 86.6%) + (0.7249 x 54.3%) was 63.2%.

Analysis of the Melbourne maternity hospital research dataset showed that the percentage of births to mothers born in an OECD country ranged from 35.9% in Dandenong Hospital to 80.9% in The Angliss Hospital. The modelled maternal CMV seroprevalence by hospital based on maternal COB classification is presented in Table 3.

**Table 3.**
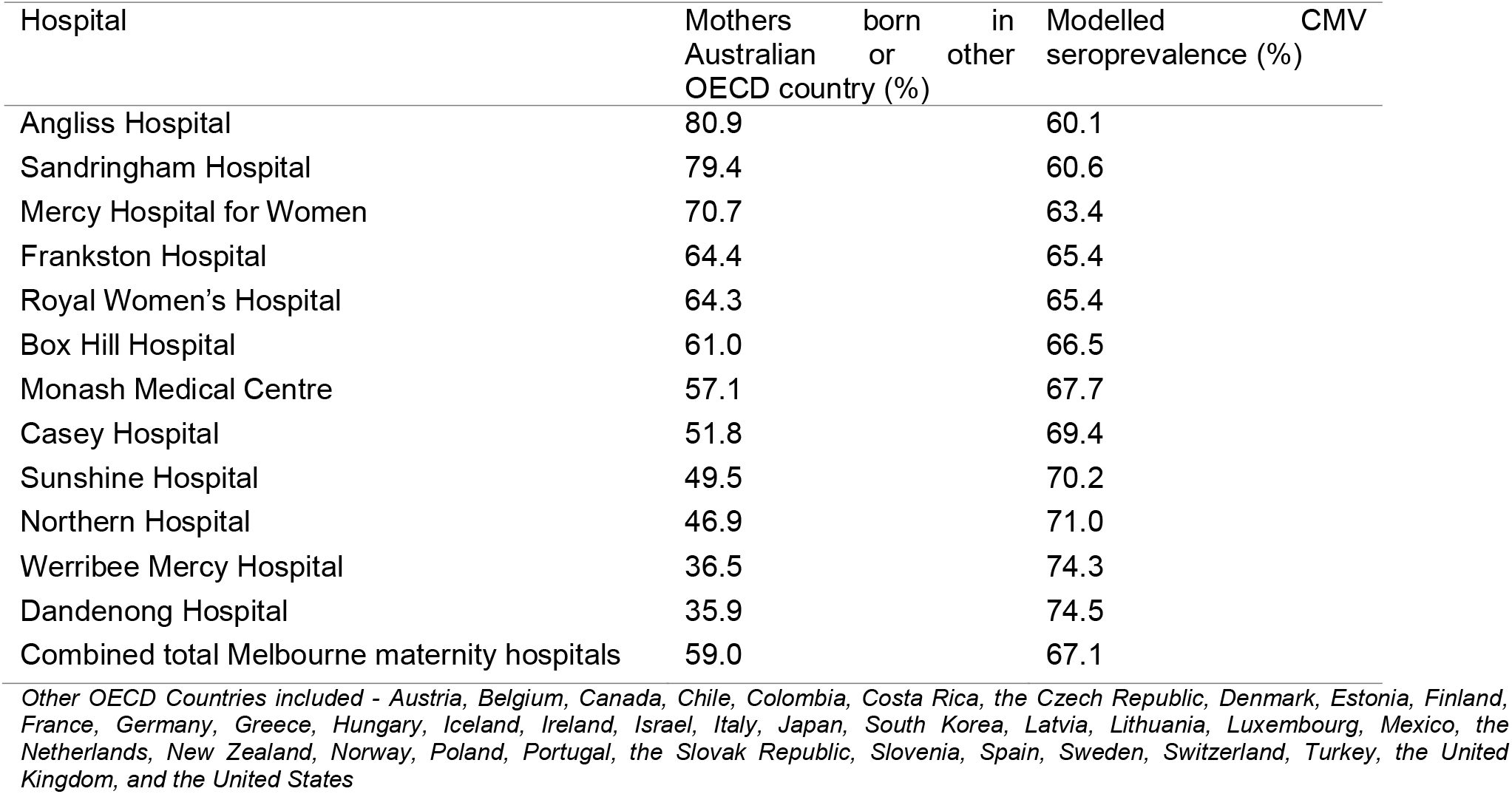
Modelled CMV seroprevalence by health service based on maternal country of birth profile.

## Discussion

This study provides contemporary Australian estimates of maternal CMV seroprevalence, derived from real-world antenatal screening data combined with national and local demographic modelling. Our results reveal significant variation in seroprevalence based on maternal country of birth and demonstrate how local population demographics can shape the risk of primary CMV infection across different healthcare settings.

The association between maternal country of birth and CMV serostatus observed in our hospital population aligns with global patterns described in the systematic review by Ssentongo et al [1]. Our estimated national seroprevalence of 63.2% is higher than earlier Australian estimates and reflects the growing diversity of our contemporary population. For comparison, Seale et al. (2006) reported a seroprevalence of 57% in a national study using remnant sera from individuals aged 1–59 years [12]. Similarly, Munro et al. (2005) found a seroprevalence of 56.8% among 600 pregnant women in an eastern Sydney [5]. Our Melbourne-based data offers more recent estimates from early pregnancy that captures demographic shifts over time and between regions.

As expected, our modelling of CMV seroprevalence across maternity services showed higher CMV seroprevalence in regions with higher migrant and refugee populations. In contrast, hospitals serving populations with a higher proportion of mothers born in Australia had lower seroprevalence. For example, approximately 40% of women giving birth at The Angliss Hospital or Sandringham Hospital were estimated to be CMV IgG negative at booking, compared with only 25% at Dandenong Hospital. We also confirmed the well-established association between higher socioeconomic status and seronegative status, though this association was of smaller magnitude than OECD country of birth.

Our results have direct implications for CMV screening at the first antenatal visit. The recently updated Commonwealth Government’s Australian Pregnancy Care Guidelines recommends clinicians: “*Offer and recommend testing early in pregnancy for cytomegalovirus to women at high risk of infection (e*.*g. with young children at home who attend childcare, or who work in childcare) using serology (cytomegalovirus-specific IgG only, not IgM)*.” CMV IgG testing combined with hygiene counselling for seronegative women at high risk of primary infection (due to personal or occupational exposure) has been shown to significantly reduce the rate of maternal CMV seroconversion from 7.6% to 1.2% [13]. While the evidence base on the effectiveness of hygiene precautions for the general antenatal population is limited [14], this advice remains the foundation of Australia’s CMV prevention strategy [15].

Importantly, hygiene advice is also recommended for seropositive women, as pre-existing CMV IgG reduces, but does not eliminate, the risk of congenital infection. Non-primary maternal CMV infections include reinfection or reactivation of latent virus. Although the fetal infection rate is much lower in non-primary maternal infections (∼1%), it can still manifest as severe disease. Indeed, on a global scale, non-primary infections are responsible for the majority of congenital CMV cases [16]. Unfortunately, no interventions have been proved to reduce fetal infections due to nonprimary maternal infection. Consequently, a pragmatic precautionary approach is to recommend hygiene counselling for all pregnant women regardless of serostatus.(Figure 2).

**Figure 2.**
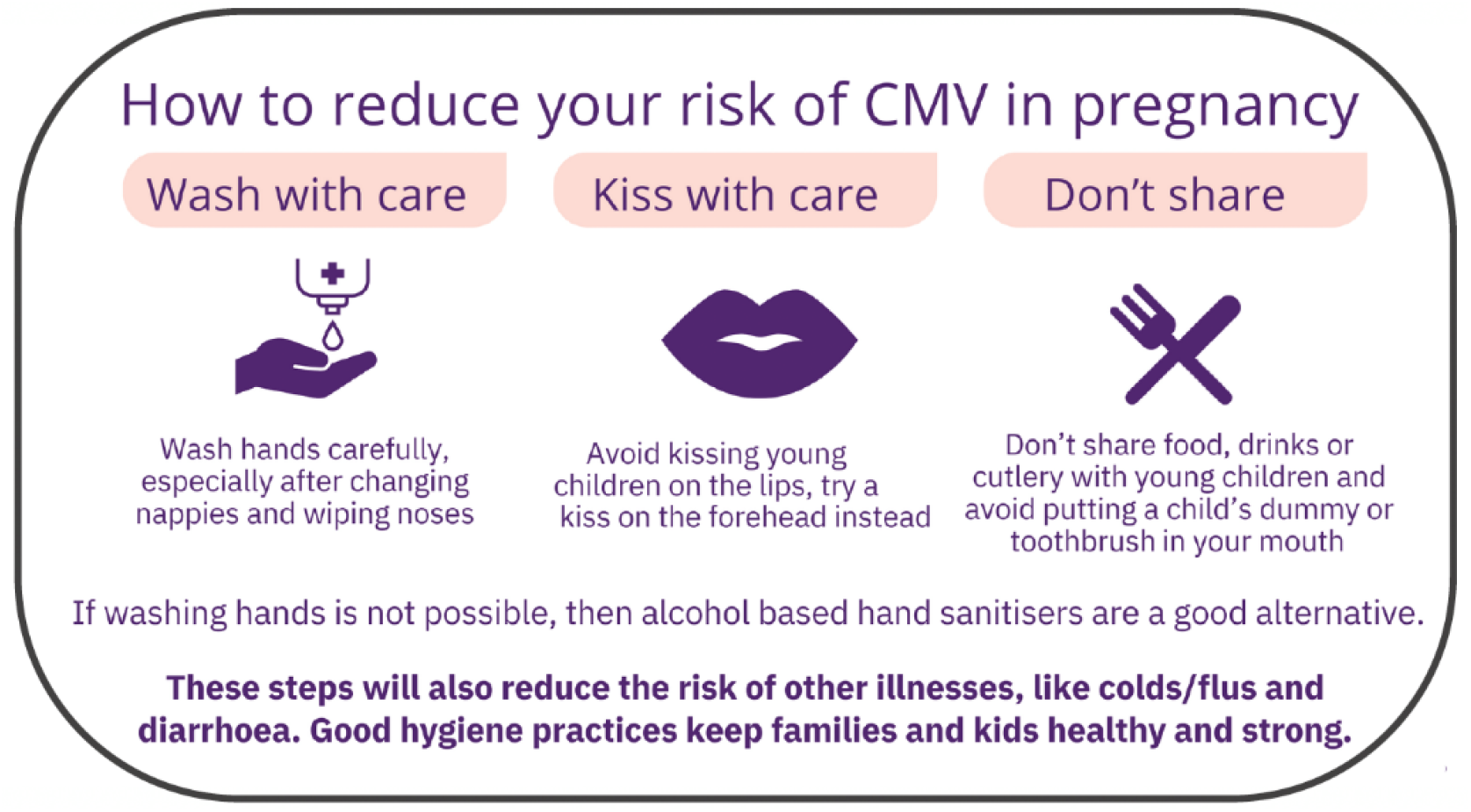
Hygiene prevention advice for pregnant women and women planning pregnancy (Image source: Cerebral Palsy Alliance)

Research has shown that Australian pregnant women find CMV hygiene messages highly acceptable. Over 90% of respondents in a recent survey evaluating a patient education video indicated that they found the information on CMV valuable with > 85% agreeing that the hygiene advice was “easy to follow”. [17]. However, less than 10% of maternity clinicians routinely provide CMV counselling at the first antenatal visit [18]. Qualitative research with Australian women underscores the importance of raising public and clinician awareness of CMV *“I found it appalling or outrageous that people in the maternity sector weren’t aware of CMV*” [19]. There are now several Australian online resources that have been proven improve clinician’s knowledge and confidence in discussing CMV prevention, including eLearning modules tailored for doctors and midwives[20, 21].

If maternal primary infection in first trimester is confirmed by seroconversion, prompt administration of high dose maternal valaciclovir has been shown to reduce the risk of fetal infection [4, 22]. In countries where seroprevalence is low, such as France and Italy, routine screening with serial serology in first trimester has been adopted to facilitate early detection and intervention with valaciclovir. A European consensus group of experts has recommended CMV serology at the first antenatal visit, with serial testing every four weeks until 14-16 weeks’ gestation for seronegative women [23]. Australia has a higher seroprevalence rate than France, Belgium or Italy, suggesting that the cost-benefit analysis of routine screening in Australia may be less favourable than in Western Europe. Health economics analyses of CMV screening strategies in Australia are needed to guide our local policy.

A key strength of this study is its real-world data on antenatal CMV screening in the early first trimester, the critical period for early implementation of CMV prevention strategies. To our knowledge, this is the only large Australian pregnancy cohort with individual-level data on maternal country of birth, parity, gestational age at testing, and sociodemographic predictors of CMV serostatus. The diversity of our cohort enhances its relevance to Australia’s multicultural population. However, several limitations should be noted. We based our CMV seroprevalence calculations on data derived from the 12.5% of women who had CMV screening in early pregnancy, which assumes they were representative of the broader antenatal population. Our hospital cohort was geographically limited to the northern Melbourne suburbs and included very few First Nations individuals. Our pragmatic approach to modelling national and Melbourne health service seroprevalence using maternal country of birth assumed that seroprevalence was similar across all countries within the OECD and non-OECD groups.

We could not model the expected seroprevalence for private hospital populations as country of birth data were not available. Further research is needed to better understand CMV serostatus in the private obstetric population, rural and remote settings, and among First Nations people.

## Conclusion

This study provides contemporary, real-world data on maternal CMV seroprevalence in Australia. Our hospital prevalence rate of 61.3% and modelled national CMV seroprevalence of 63.2% are higher than previously reported for an Australian maternity population. These findings highlight the importance of considering population diversity in CMV prevention strategies and the value of early antenatal IgG screening to identify women at risk of primary infection. These data are particularly relevant considering the international evidence supporting serial testing and antiviral intervention for maternal primary infections. Further research is needed to assess the effectiveness, acceptability, and cost-effectiveness of CMV prevention strategies in diverse settings and maternity care models.

## Data Availability

All data produced in the present study are available upon reasonable request to the authors and HREC approval

## Acknowledgements

xxxx

